# Evaluation of the Implementation of the 4C Mortality Score in United Kingdom hospitals during the second pandemic wave

**DOI:** 10.1101/2021.12.18.21268003

**Authors:** Andrew E. Blunsum, Jonathan S. Perkins, Areeb Arshad, Sukrit Bajpai, Karen Barclay-Elliott, Sanjita Brito-Mutunayagam, Rebecca Brooks, Terrence Chan, Dominic J. G. Coates, Alina Corobana, Tim Crocker-Buqué, Terry J. Evans, Jasmine Gordon-Brown, Berkin Hack, Heather Hiles, Aakash Khanijau, Salina Lalwani, Clare Leong, Kirsty MacKay, Catriona Macrae, Bryony Martin, Christopher A. Martin, Emily McKemey, Joshua Nazareth, Daniel Pan, Marcello Scopazzini, David Simons, Sophie Swinhoe, Julia Thomas, Fiona Thorburn, Sarah Walpole, Esmie Warne, Rory Wilson, Alisdair MacConnachie, Antonia Ho

**Affiliations:** Queen Elizabeth University Hospital, Glasgow; Glasgow Royal Infirmary, Glasgow; Forth Valley Royal Hospital, Larbert; Queen Elizabeth Hospital, Birmingham; University Hospital Coventry, Coventry; St. John’s Hospital, Livingston; Western General Hospital, Edinburgh; Addenbrooke’s Hospital, Cambridge; Kingston Hospital, Kingston upon Thames; University Hospital Wishaw, Wishaw; St. George’s Hospital, London; Royal Free Hospital, London; University Hospital Hairmyres, East Kilbride; Newham University Hospital, London; Royal Hallamshire Hospital, Sheffield; Warrington Hospital, Warrington; Edinburgh Royal Infirmary, Edinburgh; Lady Margaret Hospital, Millport; University Hospital Crosshouse, Kilmarnock; Arran War Memorial Hospital, Lamlash; University Hospital Ayr, Ayr; University Hospital Monklands, Airdrie; Royal Alexandra Hospital, Paisley; Leicester General Hospital, Leicester; Glenfield Hospital, Leicester; Leicester Royal Infirmary, Leicester; Department of Respiratory Sciences, University of Leicester; Victoria Hospital, Kirkcaldy; University College Hospital, London; Dumfries and Galloway Royal Infirmary, Dumfries; Galloway Community Hospital, Stranraer; Royal Victoria Infirmary, Newcastle; John Radcliffe Hospital, Oxford; Ninewells Hospital, Dundee; MRC-University of Glasgow Centre for Virus Research, Glasgow

**Author notes:** **Correspondence to:** Dr. Antonia Ho, MRC-University of Glasgow Centre for Virus Research, University of Glasgow, Glasgow, G61 1QH, **Email:**. Joint first authors (Andrew E. Blunsum, Jonathan S. Perkins).

**Keywords:** 4C Mortality Score, 4C Score, COVID-19, COVID-19 management guidance, inclusion of 4C score, documentation of 4C score

## Abstract

The 4C Mortality Score (4C Score) was designed to risk stratify hospitalised patients with COVID-19. We assessed inclusion of 4C Score in COVID-19 management guidance and its documentation in patients’ case notes in January 2021 in UK hospitals. 4C Score was included within guidance by 50% of sites, though score documentation in case notes was highly variable. Higher documentation of 4C Score was associated with score integration within admissions proformas, inclusion of 4C Score variables or link to online calculator, and management decisions. Integration of 4C Score within clinical pathways may encourage more widespread use.

## INTRODUCTION

Early identification of patients with COVID-19 at risk of poor outcome could support early decision-making regarding treatment and escalation of care (1). The 4C Mortality score (4C Score) was developed to better inform clinical decision-making in patients hospitalised with COVID-19 (2), and has been validated in COVID-19 patients hospitalised between August 2020 and February 2021 (3). This study aimed to assess if, and how, the 4C Score was incorporated into the assessment of COVID-19 patients in UK hospitals in January 2021 during the peak of the second wave (4).

## METHODS

Between 3 June and 30 September 2021, investigators from all UK hospitals were invited to take part in a cross-sectional study on COVID-19-specific management guidance at their hospital site, and the role of the 4C Score within these documents. Additionally, they were invited to undertake a retrospective case note review of adult patients presenting with community-acquired COVID-19 during a 2-week period (11-24 January 2021) at the peak of the ‘second wave’ to ascertain the proportion of patients that had a 4C Score documented in their case notes. The protocol was developed in collaboration with the UK National Infection Trainee Collaborative for Audit and Research (NITCAR) following STROBE guidance (5). Site investigators were recruited through NITCAR and local trainee networks. Data were collected on a REDCap database (Research Electronic Data Capture, Vanderbilt University, US) (6). This study was classified as service evaluation by the West of Scotland Research Ethics Committee. Caldicott Guardian approval was obtained at each participating site. Details of the study protocol are included as an online Appendix.

## RESULTS

Forty-one hospitals in England and Scotland participated in the cross-sectional study of COVID-19 specific guidance, 32 of which took part in the retrospective cohort study (Supplementary Table S1). Characteristics of participating hospitals are summarised in Table 1.

**Table 1.**
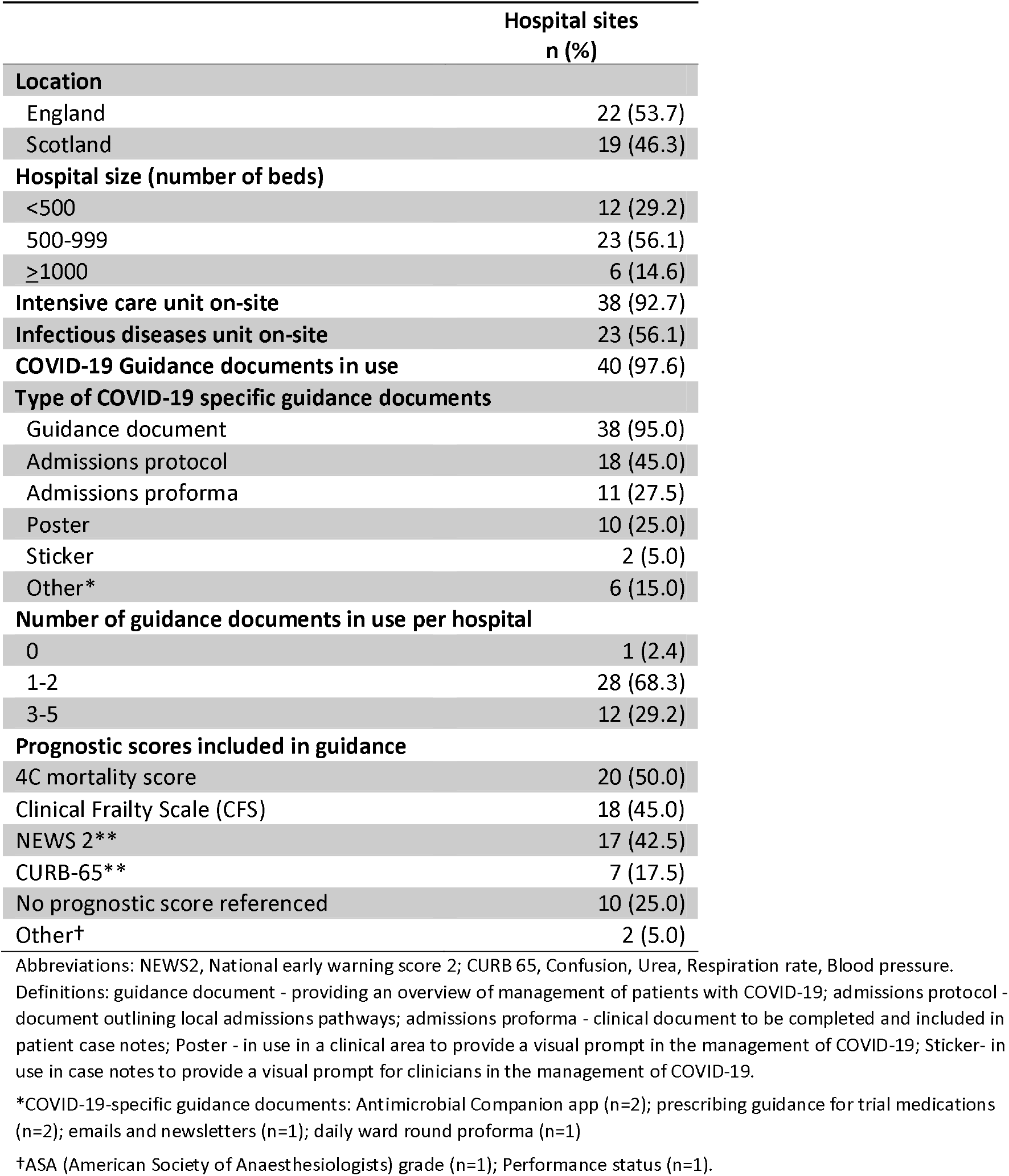
Characteristics and guidance document use in participating hospitals

|  | Hospital sites<br>n (%) |
| --- | --- |
| **Location** |  |
| England | 22 (53.7) |
| Scotland | 19 (46.3) |
| **Hospital size (number of beds)** |  |
| <500 | 12 (29.2) |
| 500-999 | 23 (56.1) |
| ≥1000 | 6 (14.6) |
| **Intensive care unit on-site** | 38 (92.7) |
| **Infectious diseases unit on-site** | 23 (56.1) |
| **COVID-19 Guidance documents in use** | 40 (97.6) |
| **Type of COVID-19 specific guidance documents** |  |
| Guidance document | 38 (95.0) |
| Admissions protocol | 18 (45.0) |
| Admissions proforma | 11 (27.5) |
| Poster | 10 (25.0) |
| Sticker | 2 (5.0) |
| Other* | 6 (15.0) |
| **Number of guidance documents in use per hospital** |  |
| 0 | 1 (2.4) |
| 1-2 | 28 (68.3) |
| 3-5 | 12 (29.2) |
| **Prognostic scores included in guidance** |  |
| 4C mortality score | 20 (50.0) |
| Clinical Frailty Scale (CFS) | 18 (45.0) |
| NEWS 2** | 17 (42.5) |
| CURB-65** | 7 (17.5) |
| No prognostic score referenced | 10 (25.0) |
| Other† | 2 (5.0) |

Abbreviations: NEWS2, National early warning score 2; CURB 65, Confusion, Urea, Respiration rate, Blood pressure.
Definitions: guidance document - providing an overview of management of patients with COVID-19; admissions protocol - document outlining local admissions pathways; admissions proforma - clinical document to be completed and included in patient case notes; Poster - in use in a clinical area to provide a visual prompt in the management of COVID-19; Sticker- in use in case notes to provide a visual prompt for clinicians in the management of COVID-19.
\*COVID-19-specific guidance documents: Antimicrobial Companion app (n=2); prescribing guidance for trial medications (n=2); emails and newsletters (n=1); daily ward round proforma (n=1)
†ASA (American Society of Anaesthesiologists) grade (n=1); Performance status (n=1).

All but one hospital (97.6%) reported the use of COVID-19-specific guidance. The most common forms of guidance were guidance documents (n=38; 95.0%) and admissions protocols (n=18; 45.0%) (Table 1; see Appendix for definitions). Twelve sites (30.0%) utilised three or more forms of COVID-19 specific guidance.

Thirty (75.0%) hospital sites referenced one or more prognostic scores in their COVID-19-specific guidance; 4C Score was the most commonly included (n=20; 50.0%), followed by Clinical Frailty Scale (CFS; n=18) and NEWS2 (n=17) (Table 1). 4C Score was utilised alongside one or more other prognostic scores at 14 sites, most commonly with CFS (n=12). Thirteen (65.0%) sites that included 4C Score within their COVID-19 guidance used it to inform management decisions, including treatment escalation (n=8), discharge (n=4), admission destination (n=3), treatment limitation (n=3) as well as remdesivir prescription (n=3).

After assessment against eligibility criteria (see Appendix), 4,123 case records from 32 hospital sites were analysed (Figure 1). 4C Score was recorded in >10% patient records in eight (25.0%) sites, ≤10% records in 10 (31.3%), and 0% in 14 (43.8%) sites (Table 2).

**Table 2.**
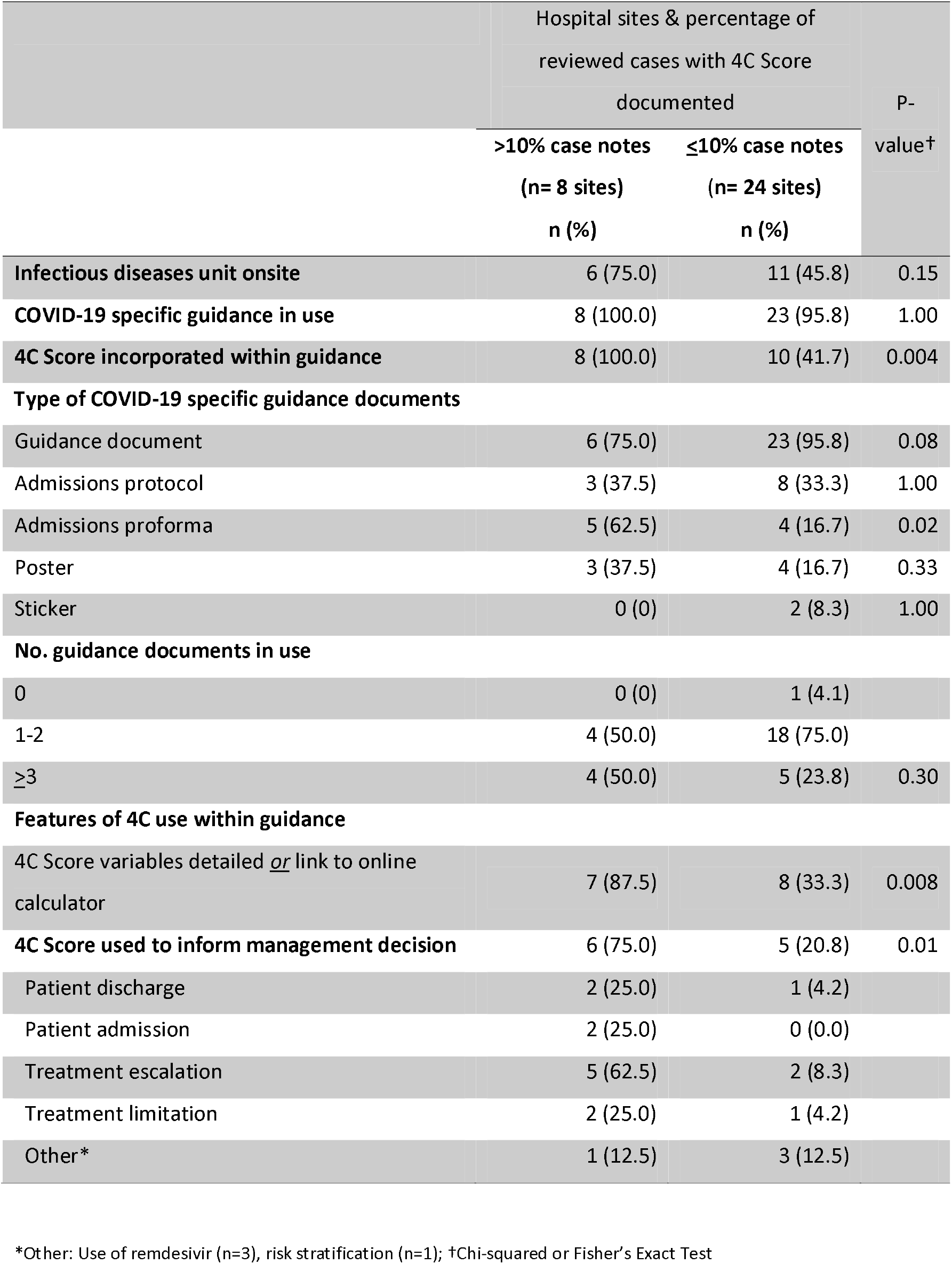
Comparison of the use of COVID-19 management guidance in hospitals that recorded 4C Score in >10 or ≤10% of patient case notes.

|  | Hospital sites & percentage of reviewed cases with 4C Score documented |  | P-value† |
| --- | --- | --- | --- |
|  | >10% case notes | ≤10% case notes |  |
|  | (n= 8 sites)<br>n (%) | (n= 24 sites)<br>n (%) |  |
| **Infectious diseases unit onsite** | 6 (75.0) | 11 (45.8) | 0.15 |
| **COVID-19 specific guidance in use** | 8 (100.0) | 23 (95.8) | 1.00 |
| **4C Score incorporated within guidance** | 8 (100.0) | 10 (41.7) | 0.004 |
| **Type of COVID-19 specific guidance documents** |  |  |  |
| Guidance document | 6 (75.0) | 23 (95.8) | 0.08 |
| Admissions protocol | 3 (37.5) | 8 (33.3) | 1.00 |
| Admissions proforma | 5 (62.5) | 4 (16.7) | 0.02 |
| Poster | 3 (37.5) | 4 (16.7) | 0.33 |
| Sticker | 0 (0) | 2 (8.3) | 1.00 |
| **No. guidance documents in use** |  |  |  |
| 0 | 0 (0) | 1 (4.1) |  |
| 1-2 | 4 (50.0) | 18 (75.0) |  |
| ≥3 | 4 (50.0) | 5 (23.8) | 0.30 |
| **Features of 4C use within guidance** |  |  |  |
| 4C Score variables detailed <u>or</u> link to online calculator | 7 (87.5) | 8 (33.3) | 0.008 |
| **4C Score used to inform management decision** | 6 (75.0) | 5 (20.8) | 0.01 |
| Patient discharge | 2 (25.0) | 1 (4.2) |  |
| Patient admission | 2 (25.0) | 0 (0.0) |  |
| Treatment escalation | 5 (62.5) | 2 (8.3) |  |
| Treatment limitation | 2 (25.0) | 1 (4.2) |  |
| Other* | 1 (12.5) | 3 (12.5) |  |

\*Other: Use of remdesivir (n=3), risk stratification (n=1); †Chi-squared or Fisher's Exact Test

**Figure 1.**
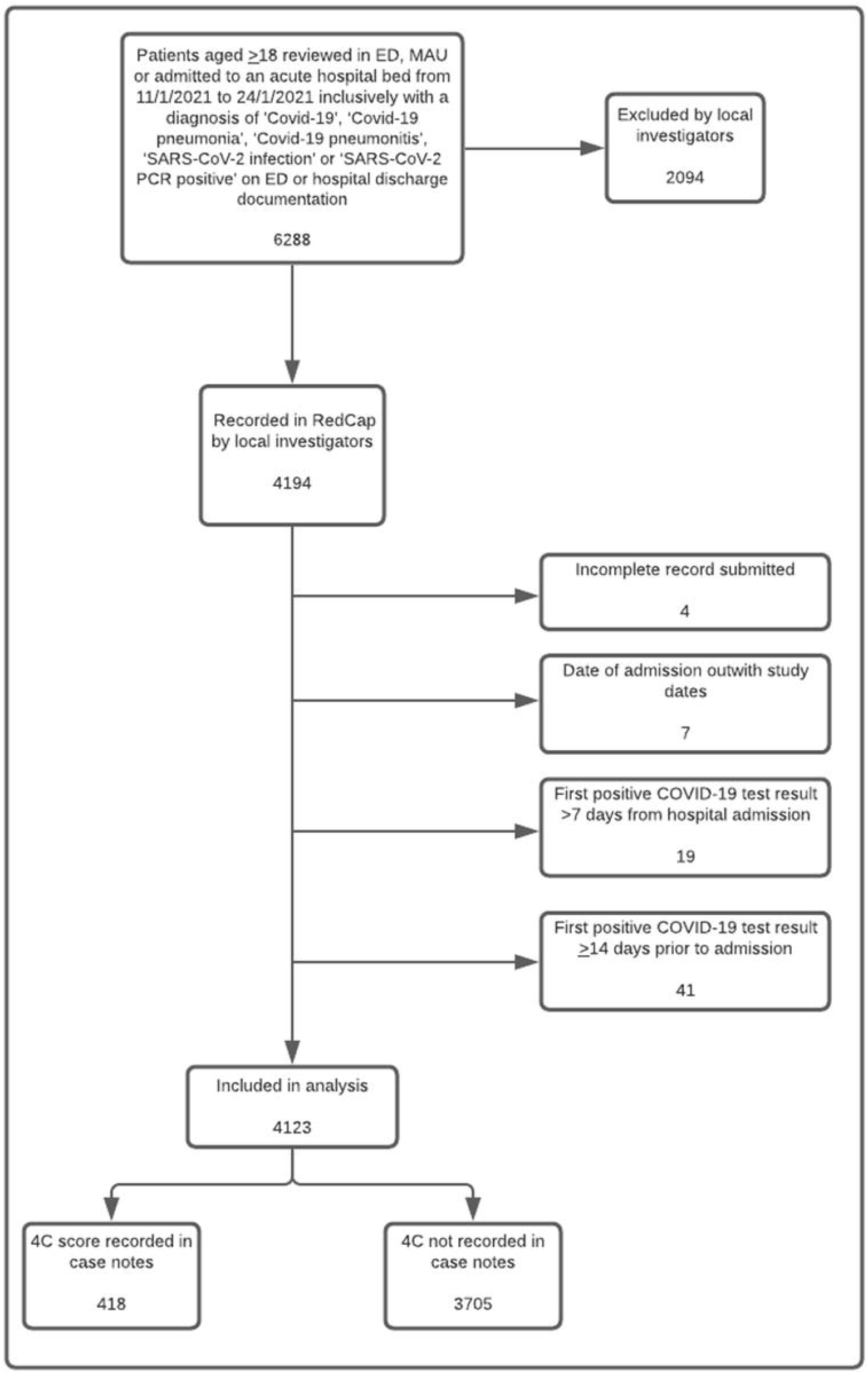
Flow chart of eligibility criteria for retrospective case note review

Overall, it was documented in the records of 418 (10.1%) patients, with wide variation in frequency of 4C Score documentation between hospital sites (median 8.2%, range 1.0-74.1% among 18 sites with 4C Score documented in ≥1 case record). The score was mostly documented by junior doctors from foundation to specialty trainees (77.5%, n=324), and predominantly in medical assessment units or medical wards (93.8%), compared to the Emergency Department (3.8%).

The eight sites that recorded 4C Score for >10% of reviewed patients were more likely to include the score within the admissions proforma (62.5% (5/8) vs. 16.7% (4/24), p= 0.02), detail the variables or incorporate an electronic link to the online calculator (87.5% (7/8) vs. 33% (8/24), p=0.008), and utilise the score to inform management recommendations (75.0% (6/8) vs. 20.8% (5/24), p=0.01).

## DISCUSSION

No prior studies have explored the use of COVID-19-specific guidance or the application of 4C Score in the assessment of COVID-19 patients. In this nationwide study, most surveyed hospitals (97.6%) had COVID-19-specific guidance documents; around two-thirds employed multiple forms of guidance. The 4C Score was the most frequently referenced prognostic score, often in conjunction with CFS. The latter is the only clinical stratification tool referenced in the NICE COVID-19 management guidance (7). The 4C Score was chiefly used to inform treatment escalation, but was also used to inform decisions on patient destination (i.e. discharge or admission location) as well as treatment limitation.

Although half of participating sites incorporated the 4C Score within guidance, documentation of the score varied substantially between sites. Audits of documentation of established mortality risk scores such as CURB-65 (for patients with community-acquired pneumonia) have also demonstrated variable recording (8,9). Nevertheless, our findings suggest that documentation of the 4C Score may be encouraged by incorporation into clinicians’ workflow through admissions proformas, detailing 4C Score variables or online calculator within guidance, in addition to linking the score to management recommendations.

This project demonstrates the capacity of national trainee networks to facilitate multi-centre studies. Collaboration with NITCAR and other trainee networks enabled the rapid recruitment of 41 UK sites and completion of >4,000 case note reviews within a short timeframe.

This study has several limitations. Although NITCAR is a national organisation, site participation was not representative, with a preponderance towards Scottish sites and no sites included from Wales or Northern Ireland. We assumed that guidance documents reported by sites between June and September 2021 were in use in January 2021 (period selected for retrospective case note review); some of these documents may not have been in use at that time or may have since been updated. We equated documentation with use of the 4C Score; this would have omitted occasions when a score was calculated but undocumented in the case notes, or conversely, decisions that were made without the value being considered.

In summary, this national study highlights that 4C Score and other clinical stratification tools were widely adopted in COVID-19-specific management guidance. Ongoing validation of the 4C Score in the context of vaccinated patients and emerging SARS-CoV-2 variants is needed. However, ensuring that the score is accessible to clinicians through integration within clinical pathways could lead to more widespread deployment.

## Supporting information

STROBE checklist

## Data Availability

All data produced in the present study are available upon reasonable request to the authors

## Acknowledgements

The authors would like to thank the following for their contribution to this study. NITCAR committee for support and assistance in facilitating this nationwide study. Vanesa Anton-Vazquez, Ali Alam, Matthew Kennedy, Imran Khan, Darver Malik, Andy Nicholson, Linda Provan, Matthew Stevens, Omolola Wilson for contributing hospital site data regarding COVID-19 specific guidance.

## Contributors

AH is guarantor and corresponding author for this work and attests that all listed authors meet authorship criteria and that no others meeting the criteria have been omitted. Contributor role taxonomy (CRediT): conceptualisation: AH, AEB, AM, JSP; data curation: AEB, JSP, AH; formal analysis AH; investigation: JSP, AA, SB, KB-E, SB-M, RB, TC, DC, AC, TC-B, JE, JG-B, BH, HH, AK, SL, CL, KM, CM, BM, CM, EM, JN, DP, LP, MS, DS, SS, JT, FT, SW, EW, RW; methodology: AH, AEB, AM, JSP; project administration: JSP, AEB, NITCAR, AH; writing – original draft: AEB, JSP, AH; writing – review and editing: AA, SB, KB-E, SB-M, RB, TC, DC, AC, TC-B, JE, JG-B, BH, HH, AK, SL, CL, KM, CM, BM, CM, EM, JN, DP, LP, MS, DS, SS JT, FT, SW, EW, RW, AM, NITCAR.

## Patient consent for publication

Not required.

