## Supplementary materials for "Evaluation of the Implementation of the 4C Mortality Score in United Kingdom hospitals during the second pandemic wave"

**Table of contents**

| **Methods** | **2** |
| --- | --- |
| **Table S1. Characteristics of contributing hospital sites** | **4** |
| **Table S2. Retrospective case note review – Patient characteristics** | **6** |
| **Figure S1. Flowchart of eligibility criteria for retrospective case note review** | **7** |
| **Figure S2. Map of hospital sites contributing to VALUE 4C study** | **8** |
| **References** | **8** |

**Methods**

**Study Design**

This study comprised an observational, cross-sectional study and a retrospective cohort study. The protocol was developed in collaboration with the UK’s National Infection Trainee Collaborative for Audit and Research (NITCAR), following STROBE guidance (1).

**Study sites and procedures**

Investigators were invited to participate via the NITCAR and UK Clinical Virology Network (UKCVN) mailing lists, infection and non-infection local trainee networks, and the British Infection Association ‘monthly digest’ newsletter. Medical education departments were contacted to promote recruitment. The study was also publicised by lead authors via social media. All acute UK hospital sites were eligible for inclusion. Medical trainees of all levels were eligible to participate on behalf of their hospital site. After completion of training, participating investigators collected data on COVID-19-specific management guidance available in the participants’ hospital sites during the study period (3^rd^ June to 30^th^ September 2021) (Part 1).

After obtaining local Caldicott Guardian approval, a retrospective cohort study (Part 2) was conducted to identify whether the 4C mortality score (4C score) was documented in the case notes of patients presenting to hospital with COVID-19 between 11^th^ to 24^th^ January 2021, a 2-week period coinciding with the peak of hospital admissions during the second pandemic wave in the UK (2). Eligible patients for case note review were identified if *all* of the following criteria were met: age 18 years and over; patient reviewed in the Emergency Department (ED), Medical Assessment Unit (MAU) or admitted to an acute hospital bed between 11^th^ January 2021 and 24^th^ January 2021; first positive COVID-19 PCR <14 days prior to presentation **or** ≤7 days of hospital admission; diagnosis of ‘COVID-19’, ‘COVID-19 pneumonia’, ‘COVID-19 pneumonitis’, ‘SARS-CoV-2 infection’ or ‘SARS-CoV-2 PCR positive’ on ED or hospital discharge documentation. Patients were excluded if *any* of the following criteria were met: first positive SARS-CoV-2 PCR ≥14 days prior to admission or review in the ED or MAU; any patient re-admitted or re-reviewed following discharge from an admission due to COVID-19; hospital-onset COVID-19 (i.e. first positive SARS-CoV-2 PCR ≥7 days following admission to an acute hospital bed. Investigators at each site approached their medical records departments to identify eligible patients for case note review, and screened cases against the eligibility criteria. Additionally, the Part 2 REDCap data collection tool incorporated prompts to ensure that each case met the eligibility criteria.

**Definitions and variables**

*Hospital characteristics*

The following hospital characteristics were recorded: number of inpatient beds, local health authority, presence of an onsite infectious diseases unit, presence of an onsite intensive care unit.

*Forms of COVID-19 specific guidance*

COVID-19-specific management guidance was defined as any form of written guidance provided to clinicians to aid in the assessment and/or management of patients with COVID-19. COVID-19-specific management guidance was categorised as follows: ‘COVID-19-specific guidance document’- defined as a document providing an overview of the management of COVID-19 patients, with or without reference to local protocols and pathways; ‘COVID-19-specific admissions protocol’- defined as a document outlining the local admissions pathway for COVID-19 patients; ‘COVID-19-specific admissions proforma’- defined as a clinical document to be completed and entered into patients’ case notes when admitting COVID-19 patients; ‘COVID-19-specific visual memory aide’- defined as a visual prompt (e.g. poster) for use in a clinical area to provide clinicians with assistance in the management of patients with COVID-19; ‘COVID-19-specific sticker’- defined as a sticker for use in patients’ case notes to provide clinicians with a prompt in the management of patients with COVID-19. A free-text option was available for investigators to detail other forms of COVID-19 specific management guidance at their hospital site.

*Scoring systems*

The prognostic scoring systems referenced by COVID- 19-specific guidance were recorded, specifically NEWS2, CURB65, qSOFA, Clinical Frailty Score (CFS) or 4C score (3–6), in addition to a free-text option to record other referenced clinical stratification tools. If the 4C score was referenced, the specific guidance document in which this had been referenced was recorded. whether the 4C score variables and corresponding values were listed, and whether a link was available to the online 4C score calculator were recorded. Furthermore, whether 4C score was used to inform management decisions such as patient discharge, admission destination, treatment escalation or limitation was also recorded.

*Case note review*

Patient demographics were recorded: age; gender; date of presentation to hospital; date of first positive SARS-CoV-2 test (earliest result available to the reviewer, either recorded in the case notes where the earliest result had been obtained prior to admission, or available via hospital laboratory systems). If the 4C score was documented in the case notes, the following data were collected: the 4C score value; whether the score had been documented by staff from ED, MAU, a medical or surgical ward, or ICU; the grade of the clinician who recorded the score. If the 4C score was not documented, no further data were recorded.

**Data sources, collection and analysis**

Data collection tools for each part of the study were designed using REDCap and piloted by the study team (7). Anonymised data were stored on a https encrypted webserver hosted by the College of Medical Veterinary and Life Sciences at the University of Glasgow. Investigators received training through video presentations accessed via Microsoft Teams.

For the retrospective case note review, case notes were reviewed by the investigator and data collected using a REDCap database (Research Electronic Data Capture, Vanderbilt University, US). Laboratory systems were reviewed for SARS-CoV-2 PCR results where necessary. No attempt was made to re-calculate the 4C score that had been documented in case notes.

**Sample size**

All UK acute hospital sites were eligible for observational, cross-sectional study of COVID-19-specific guidance. For the retrospective case note review, all patients met the inclusion criteria were included. Investigators were eligible for citable authorship after submitting a minimum dataset of 100 records for case note review. Investigators that submitted data for the cross-sectional study only were eligible for a certificate of audit participation.

**Statistical analysis**

Analysis was performed with Stata software (version 16.1). Categorical variables are summarised as frequencies and percentages. Continuous variables are presented as medians and interquartile range (IQR). We tested differences in categorical variables using χ^2^ or Fisher exact test, and differences in continuous variables using *t* or Wilcoxon rank sum test.

**Bias**

To mitigate the risk of bias towards hospital sites with onsite infectious diseases units, investigators were encouraged to recruit colleagues from other hospital sites within their health boards. Efforts were made to advertise the study and recruit investigators through non-infection trainee networks nationwide.

**Table S1. Characteristics of contributing hospital sites**

| Site Name | Onsite ID unit | Onsite ITU | No. of guidance documents used | No. of case notes reviewed for Part 2 | Percentage of cases with 4C Mortality Score recorded  (N, %) |
| --- | --- | --- | --- | --- | --- |
| Dumfries & Galloway Royal Infirmary | Yes | Yes | 2 | 81 | 60 (74.1) |
| Glenfield Hospital | No | Yes | 4 | 56 | 36 (64.3) |
| Western General Hospital | Yes | Yes | 3 | 71 | 33 (46.5) |
| University College London Hospital | Yes | Yes | 4 | 246 | 110 (44.7) |
| Newham University Hospital | Yes | Yes | 1 | 100 | 35 (35.0) |
| Royal Hallamshire Hospital | Yes | Yes | 3 | 60 | 20 (33.3) |
| Leicester Royal Infirmary | Yes | Yes | 2 | 358 | 69 (19.3) |
| Galloway Community Hospital | No | No | 1 | 24 | 3 (12.5) |
| University Hospital Monklands | Yes | Yes | 1 | 143 | 12 (8.4) |
| Ninewells Hospital | Yes | Yes | 0 | 108 | 9 (8.3) |
| Addenbrookes Hospital | Yes | Yes | 3 | 169 | 10 (5.9) |
| St John’s Hospital | No | Yes | 1 | 44 | 2 (4.5) |
| Kingston Hospital | No | Yes | 1 | 109 | 4 (3.7) |
| Queen Elizabeth University Hospital, Glasgow | Yes | Yes | 1 | 107 | 4 (3.7) |
| Glasgow Royal Infirmary | No | Yes | 2 | 208 | 6 (2.9) |
| Edinburgh Royal Infirmary | No | Yes | 3 | 96 | 2 (2.1) |
| Victoria Hospital, Fife | No | Yes | 5 | 95 | 1 (1.1) |
| Queen Elizabeth Hospital, Birmingham | No | Yes | 1 | 207 | 2 (1.0) |
| Aberdeen Royal Infirmary | Yes | Yes | 2 | n/a | n/a |
| Arran War Memorial Hospital | No | No | 1 | 1 | 0 |
| Forth Valley Royal Hospital | Yes | Yes | 2 | 62 | 0 |
| James Cook Hospital | Yes | Yes | 4 | n/a | n/a |
| John Radcliffe Hospital | Yes | Yes | 2 | 101 | 0 |
| Kettering General Hospital | No | Yes | 1 | n/a | n/a |
| Lady Margaret Hospital | No | No | 1 | n/a | n/a |
| Leicester General Hospital | No | Yes | 2 | 12 | 0 |
| North Manchester General Hospital | Yes | Yes | 2 | n/a | n/a |
| Queen Elizabeth Hospital, London | No | Yes | 2 | n/a | n/a |
| Royal Alexandra Hospital | No | Yes | 2 | 102 | 0 |
| Royal Free Hospital | Yes | Yes | 2 | 375 | 0 |
| Royal Liverpool University Hospital | Yes | Yes | 3 | n/a | n/a |
| Royal Sussex County Hospital | Yes | Yes | 2 | n/a | n/a |
| Royal Victoria Infirmary | Yes | Yes | 1 | 100 | 0 |
| St George’s Hospital | Yes | Yes | 3 | 262 | 0 |
| St Helier Hospital | Yes | Yes | 4 | n/a | n/a |
| University Hospital Ayr | No | Yes | 1 | 36 | 0 |
| University Hospital Coventry | Yes | Yes | 1 | 265 | 0 |
| University Hospital Crosshouse | Yes | Yes | 1 | 89 | 0 |
| University Hospital Hairmyres | No | Yes | 2 | 109 | 0 |
| University Hospital Wishaw | No | Yes | 4 | 125 | 0 |
| Warrington Hospital | No | Yes | 2 | 203 | 0 |

| Table S2. Patient characteristics, retrospective case note review | | |
| --- | --- | --- |
|  | **4C Mortality Score recorded in case notes** | |
|  | **Yes**  **N=418**  **N (%)** | **No**  **N=3705**  **N (%)** |
| Age (years) - median (IQR) | 64.5 (55-77) | 65 (52-79) |
| Gender - male | 253 (60.5) | 1899 (51.3) |
| Risk Category (4C score) |  |  |
| Low (0-3) | 41 (9.8) | - |
| Intermediate (4-8) | 136 (32.5) | - |
| High (9-14) | 184 (44.0) | - |
| Very high (≥15) | 57 (13.6) | - |
| Clinician documenting 4C score |  |  |
| Foundation trainee | 51 (12.2) | - |
| ANP/ PA | 3 (0.7) | - |
| SHO | 168 (40.2) | - |
| Specialty trainee | 105 (25.1) | - |
| Consultant | 82 (19.6) | - |
| Unknown | 9 (2.2) | - |
| Location of 4C score documentation |  |  |
| Emergency Department | 16 (3.8) | - |
| Medical Assessment Unit | 275 (65.8) | - |
| Medical ward | 117 (28.0) | - |
| Intensive Care Unit | 7 (1.7) | - |
| Unknown | 3 (0.7) | - |

Abbreviations: ID, Infectious Diseases; ANP, Advanced Nurse Practitioners; PA, Physicians’ Assistants; SHO, Senior House Officer, also catch-all term for Core trainee, clinical fellow, and locum doctor above the level of Foundation year 2 but below the level of Specialty trainee/ registrar.

**Figure S1.** Flow chart of eligibility criteria for retrospective case note review

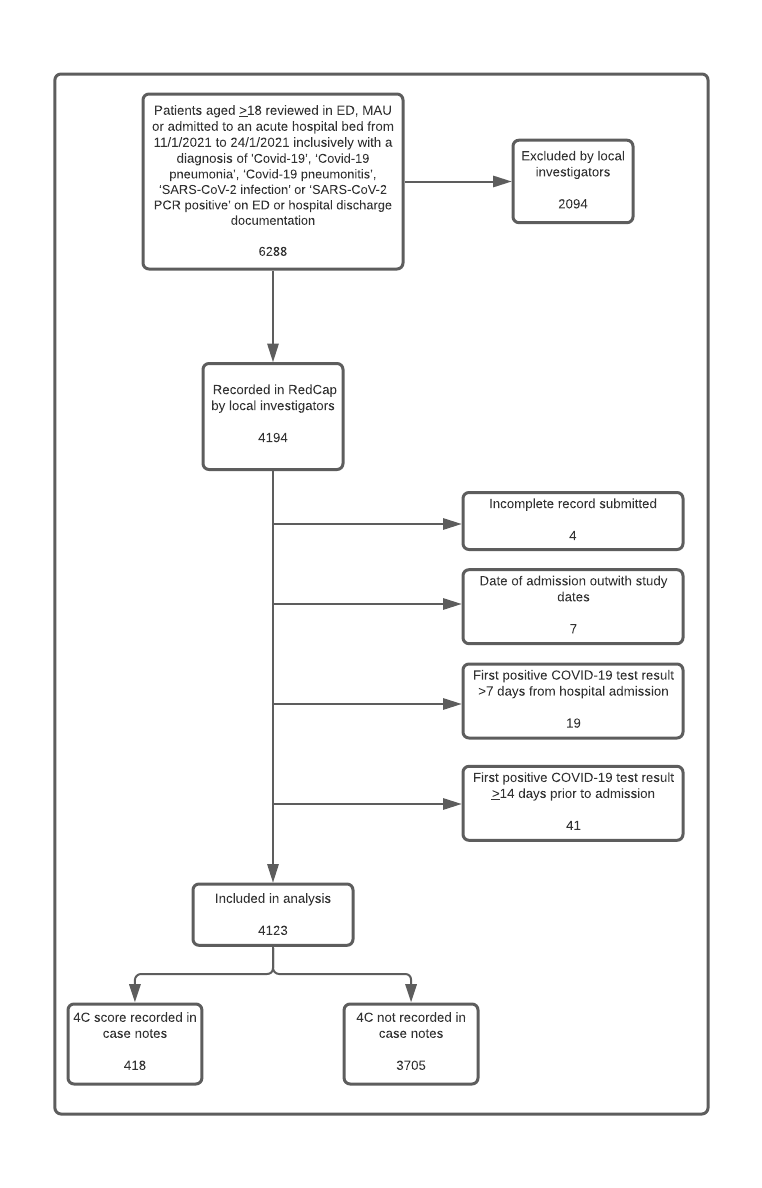

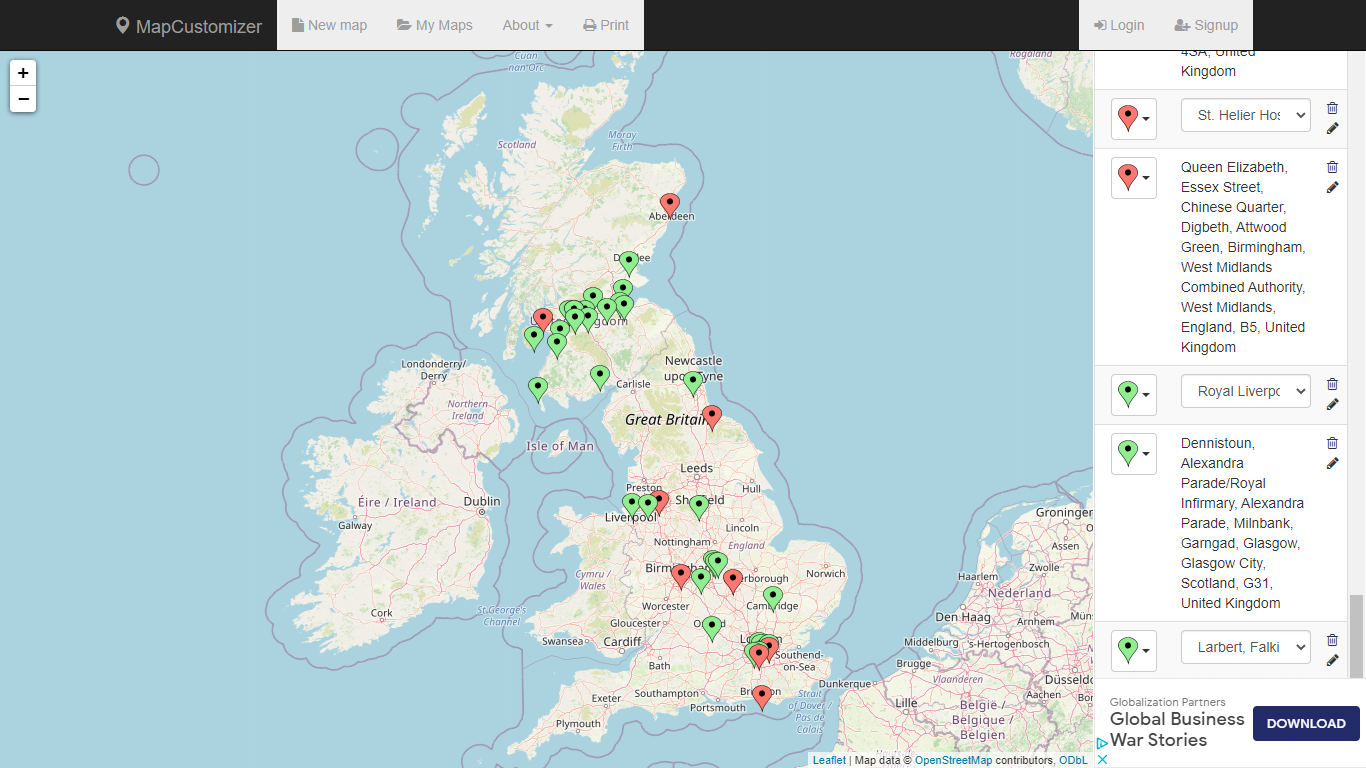
**Figure S2. Map of hospital sites contributing to VALUE 4C study**

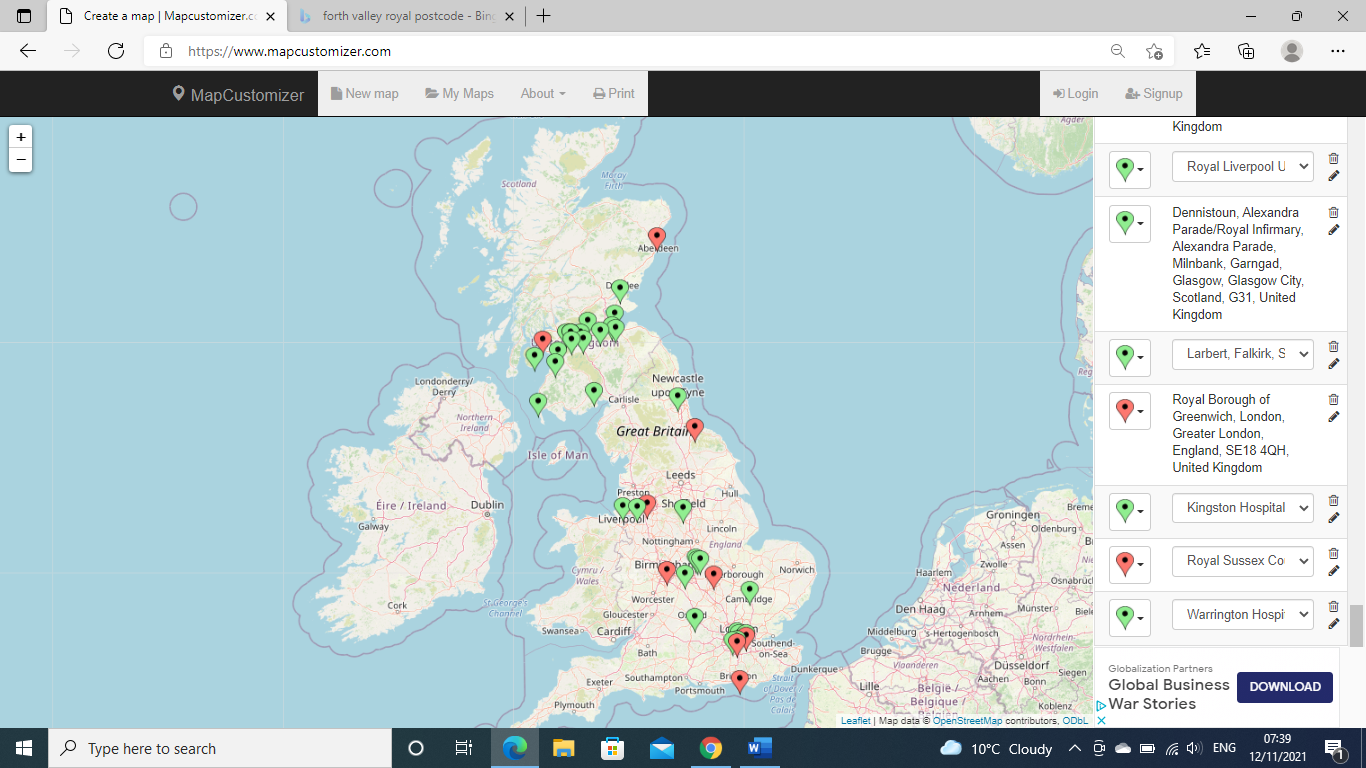

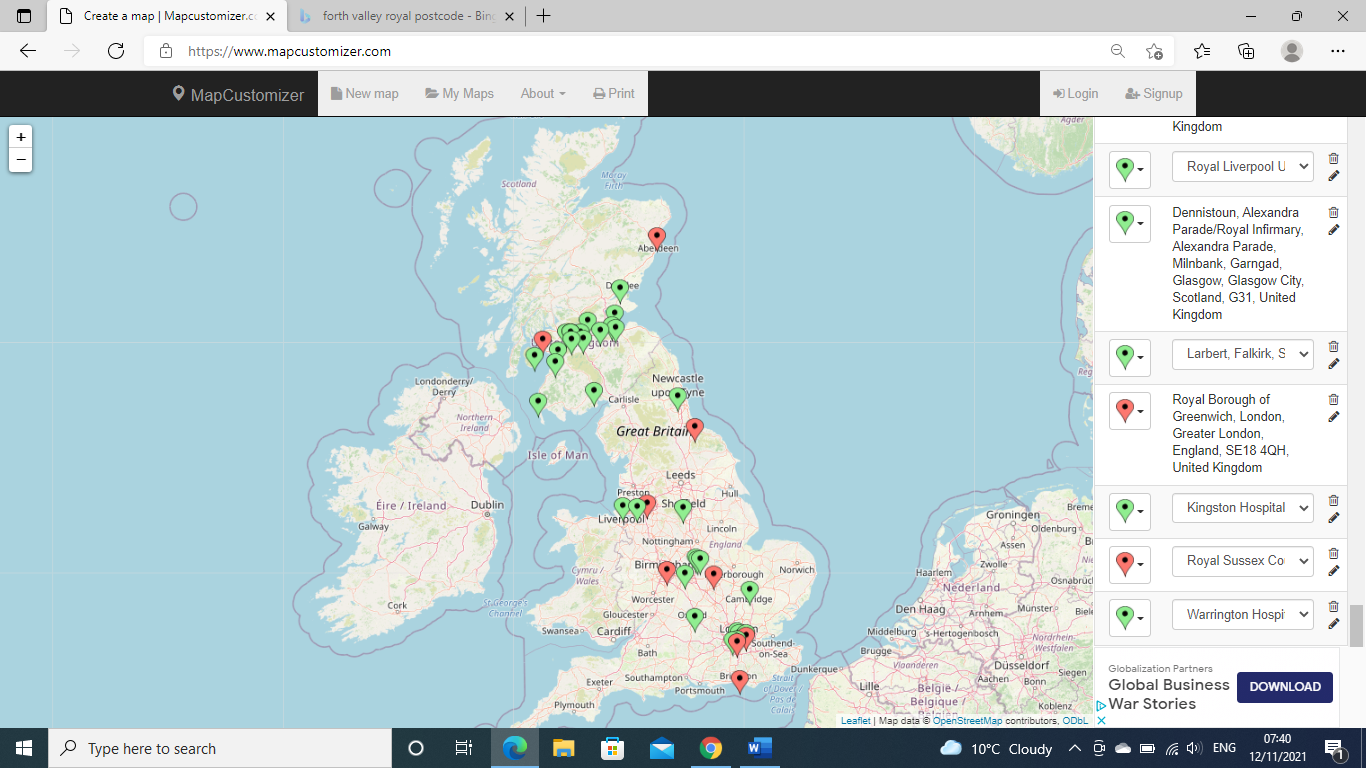

Contributed to Part 1 data only

Contributed to Part 1 and Part 2 data
