## Supplementary material for "Evaluation of the Implementation of the 4C Mortality Score in United Kingdom hospitals during the second pandemic wave": STROBE checklist

**STROBE Statement—checklist of items that should be included in reports of observational studies**

|  | Item No | Recommendation | Page  No |
| --- | --- | --- | --- |
| **Title and abstract** | 1 | (*a*) Indicate the study’s design with a commonly used term in the title or the abstract | Manuscript p2 |
|  |  | (*b*) Provide in the abstract an informative and balanced summary of what was done and what was found | Manuscript p2 |
| Introduction | | | |
| Background/rationale | 2 | Explain the scientific background and rationale for the investigation being reported | Manuscript p3 |
| Objectives | 3 | State specific objectives, including any prespecified hypotheses | Manuscript p3 |
| Methods | | | |
| Study design | 4 | Present key elements of study design early in the paper | Manuscript p3 |
| Setting | 5 | Describe the setting, locations, and relevant dates, including periods of recruitment, exposure, follow-up, and data collection | Manuscript p3, Supplement p2 |
| Participants | 6 | (*a*) *Cohort study*—Give the eligibility criteria, and the sources and methods of selection of participants. Describe methods of follow-up  *Case-control study*—Give the eligibility criteria, and the sources and methods of case ascertainment and control selection. Give the rationale for the choice of cases and controls  *Cross-sectional study*—Give the eligibility criteria, and the sources and methods of selection of participants | Manuscript p3, Supplement p2-3 |
|  |  | (*b*) *Cohort study*—For matched studies, give matching criteria and number of exposed and unexposed  *Case-control study*—For matched studies, give matching criteria and the number of controls per case |  |
| Variables | 7 | Clearly define all outcomes, exposures, predictors, potential confounders, and effect modifiers. Give diagnostic criteria, if applicable | Supplement p3 |
| Data sources/ measurement | 8* | For each variable of interest, give sources of data and details of methods of assessment (measurement). Describe comparability of assessment methods if there is more than one group | Manuscript p3 |
| Bias | 9 | Describe any efforts to address potential sources of bias | Supplement p4 |
| Study size | 10 | Explain how the study size was arrived at | Manuscript p3, Supplement p3 |
| Quantitative variables | 11 | Explain how quantitative variables were handled in the analyses. If applicable, describe which groupings were chosen and why | Supplement p3 |
| Statistical methods | 12 | (*a*) Describe all statistical methods, including those used to control for confounding | Supplement p3 |
|  |  | (*b*) Describe any methods used to examine subgroups and interactions | n/a |
|  |  | (*c*) Explain how missing data were addressed | n/a |
|  |  | (*d*) *Cohort study*—If applicable, explain how loss to follow-up was addressed  *Case-control study*—If applicable, explain how matching of cases and controls was addressed  *Cross-sectional study*—If applicable, describe analytical methods taking account of sampling strategy | n/a |
|  |  | (*e*) Describe any sensitivity analyses | n/a |

Continued on next page

| Results | | | |
| --- | --- | --- | --- |
| Participants | 13* | (a) Report numbers of individuals at each stage of study—eg numbers potentially eligible, examined for eligibility, confirmed eligible, included in the study, completing follow-up, and analysed | Manuscript p3 |
|  |  | (b) Give reasons for non-participation at each stage | Manuscript p3 |
|  |  | (c) Consider use of a flow diagram | Manuscript p9 (Figure 1) |
| Descriptive data | 14* | (a) Give characteristics of study participants (eg demographic, clinical, social) and information on exposures and potential confounders | Manuscript p4-6, Supplement p4-5 |
|  |  | (b) Indicate number of participants with missing data for each variable of interest | Figure 1 |
|  |  | (c) *Cohort study*—Summarise follow-up time (eg, average and total amount) | Manuscript p3 |
| Outcome data | 15* | *Cohort study*—Report numbers of outcome events or summary measures over time | Manuscript p5-7 |
|  |  | *Case-control study—*Report numbers in each exposure category, or summary measures of exposure | n/a |
|  |  | *Cross-sectional study—*Report numbers of outcome events or summary measures | Manuscript p3-5 |
| Main results | 16 | (*a*) Give unadjusted estimates and, if applicable, confounder-adjusted estimates and their precision (eg, 95% confidence interval). Make clear which confounders were adjusted for and why they were included | n/a |
|  |  | (*b*) Report category boundaries when continuous variables were categorized | n/a |
|  |  | (*c*) If relevant, consider translating estimates of relative risk into absolute risk for a meaningful time period | n/a |
| Other analyses | 17 | Report other analyses done—eg analyses of subgroups and interactions, and sensitivity analyses | Manuscript p7 |
| Discussion | | | |
| Key results | 18 | Summarise key results with reference to study objectives | Manuscript p7-8 |
| Limitations | 19 | Discuss limitations of the study, taking into account sources of potential bias or imprecision. Discuss both direction and magnitude of any potential bias | Manuscript p7 |
| Interpretation | 20 | Give a cautious overall interpretation of results considering objectives, limitations, multiplicity of analyses, results from similar studies, and other relevant evidence | Manuscript p7-8 |
| Generalisability | 21 | Discuss the generalisability (external validity) of the study results | Manuscript p7-8 |
| Other information | | | |
| Funding | 22 | Give the source of funding and the role of the funders for the present study and, if applicable, for the original study on which the present article is based | Manuscript p9 |

*Give information separately for cases and controls in case-control studies and, if applicable, for exposed and unexposed groups in cohort and cross-sectional studies.

**Note:** An Explanation and Elaboration article discusses each checklist item and gives methodological background and published examples of transparent reporting. The STROBE checklist is best used in conjunction with this article (freely available on the Web sites of PLoS Medicine at http://www.plosmedicine.org/, Annals of Internal Medicine at http://www.annals.org/, and Epidemiology at http://www.epidem.com/). Information on the STROBE Initiative is available at www.strobe-statement.org.
